# Does frequency or diversity of leisure activity matter more for epigenetic ageing? Analyses of arts engagement and physical activity

**DOI:** 10.1101/2024.11.01.24316559

**Authors:** Daisy Fancourt, Lehané Masebo, Saoirse Finn, Hei Wan Mak, Feifei Bu

## Abstract

Over the past decade, ageing clocks have become widely adopted as important tools for understanding biological ageing and have been redefining notions of “pro-longevity” lifestyles. However, this work is still at an early stage. Some leisure activities, such as arts and cultural engagement (ACEng) have never been studied at all, while others such as physical activity (PA) have only received scant attention. In particular, there is little understanding of whether frequency of engagement or diversity (which provides access to more active ingredients) is more important. This study used data from 3,354 adults in the UK Household Longitudinal Study - a large, nationally-representative cohort study, which includes seven derived epigenetic clocks. We used a doubly robust estimation using the inverse-probability-weighted regression adjustment estimator adjusted for demographic, socioeconomic, behavioural and health covariates, data collection gaps and technical covariates of epigenetic clocks. ACEng and PA were related to slower biological ageing in the PhenoAge, DunedinPoAm and DunedinPACE clocks, although not to the other measured clocks PA (Lin, Horvath2018, Horvath2013 and Hannum), with comparable effect sizes between ACEng. For ACEng, diversity and frequency of engagement were related to DunedinPoAm and DunedinPACE, while for PhenoAge, there was a slightly clearer relationship for frequency than diversity. For PA, higher levels of frequency, diversity, and activeness were related to DunedinPoAm and DunedinPACE, while only the highest diversity and activeness were related to PhenoAge. These results were all stronger amongst middle-aged and older adults. Our findings support future exploration of whether lifestyle changes can slow epigenetic ageing.

**Translational significance:** Population ageing poses a major global challenge, underscoring the urgency of investigating how modifiable lifestyle factors influence ageing process and healthy lifespan. This is the first study to link arts and cultural engagement with epigenetic ageing. Analysing data from a nationally representative cohort study, we found comparable effect sizes between arts and cultural engagement and physical activities. These findings extend existing research beyond specific health conditions and mortality, advancing our understanding of potential impacts of leisure activities on biological hallmarks of ageing that underpin health and diseases. Our study provides important implications for developing evidence-based interventions to delay biological ageing and promote healthy longevity.

## Introduction

With ageing populations becoming a global phenomenon, how to support not just a longer lifespan but also an increased “healthspan” is becoming a key question for both individuals and health services (The Lancet Public Health, 2017). Helping adults stay free from disease, maintain functional independence, and reduce the need for health services are key priorities for governments internationally (Beard & Bloom, 2015). Over the past two decades, theoretical and technological advances within molecular biology have identified a series of fundamental biological hallmarks of ageing, including various molecular, cellular, and systemic processes underpinning health and disease (López-Otín et al., 2023). One of these is epigenetic alterations, including alterations in DNA methylation (DNAm) patterns, aberrant chromatin remodelling, abnormal post-translational modification of histones, and deregulated function of non-coding RNAs. Environmental stress accumulated over the lifetime disrupts epigenetic profiles, and this contributes to the ageing phenotype by promoting instability, carcinogenesis and cardiovascular pathologies (Pagiatakis et al., 2021; Zhang et al., 2020). In recent years, there have been major developments in biohorology: the use of ageing “clocks” that are built from mapping patterns of DNAm across sparse but informative sets of specific CpG (cytosine-phospho-guanine) sites on the genome (Horvath & Raj, 2018). These epigenetic clocks are used to identify discrepancies between chronological vs biological age (i.e. accelerated vs decelerated ageing). While first-generation clocks were based on chronological age (e.g. Hannum, Horvath2013, Horvath2018, Lin), second-generation clocks have been developed based on phenotypic age (e.g. PhenoAge), and lifespan (e.g. GrimAge) and third-generation clocks are based on the pace of ageing (e.g. DunedinPoAm, DunedinPACE).

Ageing clocks are not without controversy: there is no gold standard for measuring epigenetic biological ageing. Ageing-related biological changes may be correlates rather than causes of ageing, the relationship between clocks and disease pathology is still in its infancy, and there is a current proliferation of clocks (Bell et al., 2019). Nonetheless, over the past decade, ageing clocks have become widely adopted as important tools for understanding biological ageing and have been redefining notions of “pro-longevity” lifestyles(Galkin et al., 2023). So, exploring ageing clocks alongside other biological approaches could provide important complementary insight into the molecular underpinnings of health. As part of this, there is increasing interest in finding modifiable lifestyle factors that might have “anti-ageing” effects. Avoiding smoking and binge drinking, maintaining a healthy body weight, adopting a Mediterranean diet, reducing stress, and engaging in meditation have all been demonstrated through combinations of experimental and epidemiological studies to reduce epigenetic age (Galkin et al., 2023).

However, this work is still at an early stage. Some leisure activities have never been studied at all. Arts and cultural engagement (ACEng) is increasingly recognised as a health behaviour in its own right, comprising diverse ‘active ingredients’ that are beneficial to health (e.g. social interaction, cognitive stimulation, multi-sensory stimulation, creativity, etc.) and activating complex psychological, biological, social and behavioural mechanisms of action that relate to mental and physical health outcomes (Fancourt et al., 2021; Fancourt & Finn, 2019). Experimental studies have already demonstrated that ACEng can affect gene regulation. For example, compared to relaxing, listening to music upregulates genes involved in processes such as dopamine secretion, enhanced synaptic function, and neurogenesis, alongside upregulating specific RNA proteins (microRNAs) that repress inflammatory cytokines and support neuronal and synaptic plasticity (Kanduri, Kuusi, et al., 2015; Kanduri, Raijas, et al., 2015; Nair et al., 2019, 2021). Music has also been demonstrated to be more effective than other activities like reading newspapers for reversing stress signatures in gene expression following laboratory-induced stressors (Bittman et al., 2005). However, there are no epidemiological studies of ACEng and epigenetic ageing to date.

Even more prominent health-promoting leisure activities, like exercise, have only received scant attention. Experimentally, physical activity (PA) has been demonstrated to cause DNAm changes. For example, people with a lifelong history of PA display lower DNAm levels on gene promoters in muscle tissue (Sailani et al., 2019), and interventions that increase PA reduce epigenetic mutation load (i.e. the total number of stochastic epigenetic mutations or outlier methylation patterns at CpG sites), which has been proposed as a complementary DNAm-based biomarker of healthy ageing (Fiorito et al., 2021). Observationally, hardly any studies have been conducted looking at physical activity and combined CpG sites within epigenetic clocks. Some very small studies (n<250) have reported null findings when relating PA to individual epigenetic clocks (Fiorito et al., 2021; Sillanpää et al., 2019). Other studies have reported associations between PA and aspects of physical performance (e.g. grip strength and jumping height) and several epigenetic clocks, including PhenoAge, FitAge and GrimAge (Jokai et al., 2023; Noroozi et al., 2024). However, these studies have failed to consider important confounders like socioeconomic position (SEP), smoking, BMI or blood cell compositions. Recently, analyses of larger cohort studies have shown more promising results. An analysis of adults in the Rhineland Study (n=3,567) found that accelerometer-derived step count and both volume and intensity of physical activity were related to lower GrimAge and PhenoAge acceleration, but not Hannum or Horvath2013 (Fox et al., 2023). And the US Sister Study (n=2,758) found that after adjusting for covariates, hours per week of leisure-time physical activity was only related to GrimAge but had no associations with Hannum, Horvath2013, or PhenoAge (Kresovich et al., 2021).

However, some key challenges remain with the existing literature. First, studies have only focused on a limited number of ageing clocks. Each ageing clock defines biological age in its own way using a distinct set of CpG sites, different primary domains (tissue, health conditions, age range), different algorithms (target definition, machine learning model), and different target variables (chronological age, phenotypic age, time-to-death, pace of ageing,etc.). Consequently, the associations between leisure activity and biological age understandably vary between clocks, meaning outcome-wide approaches using multiple ageing clocks are important for drawing broader conclusions (Bell et al., 2019). Second, *how* leisure is conceptualised and defined is underexplored. While frequency is a standard metric of people’s engagement, a variety of engagement may also be important. Variety provides people with greater opportunity to access different ‘active ingredients’ of leisure - i.e. different patterns of cognitive, physical and social stimuli, which may have different mechanistic pathways to biological ageing (Warran et al., 2022). Additionally, social identity theory posits that engagement with multiple groups provides more diverse social identities, which can be crucial to psychological processes of stress-buffering, coping and resilience (Hogg, 2016). Even when leisure is not overtly social, it can bring personal identities as being part of a collective who does that activity (e.g. “runner” or “artist”). Third, it is crucial to take account of diverse confounding factors. Previous analyses have largely relied on conditioning on confounders via simple regression adjustment. However, this leaves the potential for residual confounding imbalance. More sophisticated statistical approaches like doubly robust estimation offer new opportunities for improving causal inference. Therefore, this study used data from a large, nationally representative cohort study involving rich phenotyping of ACEng and PA, seven different epigenetic clocks, and a doubly robust statistical approach to provide new insight into the relationship between leisure and epigenetic ageing.

## Materials and Methods

### Data

Understanding Society, the UK Household Longitudinal Study (UKHLS), is a nationally representative panel survey of members of 40,000 private households in the UK. It was launched in 2009, with participants being followed up annually. Detailed information on the sampling strategy can be found in the sampling design report (Lynn, 2009). We used data from the DNAm subsample. Between 2010 and 2012 (waves 2 and 3), blood samples were collected from adult participants during nurse visits. DNAm profiling was conducted from blood samples of 3,654 eligible individuals of white European ancestry who had consented to blood sampling and genetic analysis (wave 2: n=1,425, wave 3: n=2,229). Over 850,000 methylation sites across the genome were measured using the Illumina Methylation EPIC BeadChip. Data were pre-processed via quality control procedures, including outlier removal, filtering poor-quality probes and quantile normalisation (Bao et al., 2022). Exposures were obtained from the wave 2 adult survey which included a special module on leisure activities. After excluding participants with missing data and outliers in outcome measures (3 standard deviations (SD) from the mean), we had an analytical sample of 3,354 (Figure S1 in the Supplement).

### Measures

#### Outcomes

UKHLS contains seven epigenetic clocks constructed from the DNAm data across three generations. First-generation clocks are trained exclusively on chronological age and include the single-tissue Hannum clock, Horvath2013 (estimated from multiple tissues/cells) (Horvath, 2013), Horvath skin & blood (Horvath2018) clock (another multi-tissue clock with improved accuracy on cultured cells) (Horvath et al., 2018), and Lin clock (based on DNAm profiles of 25 cancer types) (Lin & Wagner, 2015).

The second-generation clocks are trained on a composite measure of mortality and disease morbidity alongside chronological age. The one available in UKHLS is the PhenoAge clock, which is based on clinical biomarkers of phenotypic age (Levine et al., 2018).

The third-generation clocks are designed to quantify paces of biological ageing rather than static status. The DunedinPoAm clock is considered the first of the third-generation clocks (Crimmins et al., 2024). It is trained on a composite measure of longitudinal changes over time in 18 biomarkers of blood chemistry and organ systems (Belsky et al., 2020). The DunedinPACE clock is an updated version of DunedinPoAm with longer follow-ups and more reliable DNA methylation probes (Belsky et al., 2022). While the first- and second-generation clocks above were measured in years, DunedinPoAm and DunedinPACE were measured in rates of biological age per chronological year.

#### Exposures

ACEng was measured by asking if participants had done anything in four sets of activities in the last 12 months (yes or no): 1) participatory arts (e.g. singing, dancing, painting, photographing, crafting), 2) receptive arts (e.g. attending art exhibitions/events), 3) visiting heritage sites (e.g. historic parks, historic buildings, monuments), 4) other cultural activities (e.g. going to museums, libraries or archives). Frequency of engagement was derived by using the highest frequency across the four sets of activities, recoded, into four categories: once or twice yearly or less, three or four times yearly, monthly, weekly. We also derived an ACEng diversity measure by counting the number of activities and splitting this into quartiles: low (0-2), medium (3-6), high (7-10) and very high (11+).

PA was measured by a list of sporting activities, including vigorous (e.g. running, swimming, boxing, cycling, football), moderate (e.g. skiing, racquet sports, angling/fishing, yoga/Pilates if age≥64) and mild (e.g. rambling, snooker, yoga/Pilates if age<65) activities. PA frequency was derived by using the highest frequency between vigorous/moderate and mild activities, coded as no, <monthly, monthly, weekly. A PA diversity measure was derived by counting the number of activities, recoded into four categories: none, one, two/three, four or more. For sensitivity analyses, we also considered a self-rated PA activeness measure on a scale of 0 to 10, which was recoded into five categories based on the distribution of the original variable: not active (0), low (1-2), medium (3-4), high (5-6), and very high (7-10).

#### Covariates

We considered a range of demographic and socioeconomic covariates, including age (range 16 to 90), age-squared, sex (female, male), marital status (single, married/cohabitating, separated/divorced/widowed), living with children (yes, no), living area (rural, urban), education (no qualification, GCSE or below, A level or above, degree or above), household income quintiles, employment status (employed, other), and area deprivation quintiles. Also accounted for are behavioural and health factors, including smoking (never smoker, ex-smoker, current smoker), drinking frequency (on a scale of 1-almost every day to 8-not at all in the last 12 months) and self-reported long-standing physical or mental impairment, illness or disability (yes, no). It is also essential to control for the gap between data collection dates between exposures and outcomes, given the blood samples of 2,229 participants were collected in wave 3. In addition, we also adjusted for a set of technical covariates of various cell composition estimates (CD8-T cells, CD4-T cells, Natural Killer cells, B cells, monocytes and granulocytes) (Bao et al., 2022). Finally, we considered Body Mass Index (BMI), which arguably could be on the causal pathway. Unlike other covariates measured at wave 2, BMI was measured during nurse visits across waves 2 and 3. It was coded into three categories: below 25, 25 and below 30, 30 and above.

### Statistical analysis

Data were analysed using doubly robust estimation using the inverse-probability-weighted regression adjustment (IPWRA) estimator. This method involves building two models to account for non-random treatment assignment: i) a regression adjustment model for the outcome and ii) a treatment-assignment model for the exposure. It uses weighted regression coefficients to compute averages of treatment-level predicted outcomes, where the weights are the estimated inverse probabilities obtained from the treatment-assignment model. IPWRA has the double-robust property: it only requires the outcome model or the treatment-assignment model to be correctly specified, not both (Wooldridge, 2010). Both models controlled for demographic, socioeconomic, behavioural and health covariates described above. The outcome model additionally controlled for data collection gaps and technical covariates of epigenetic clocks. The IPWRA model was fitted separately for each exposure (frequency, ACEng diversity, ACEng, PA frequency, and PA diversity). We conducted sensitivity analyses that (i) considered self-reported levels of activeness in PA, (ii) additionally controlled for BMI, and (iii) restricted the sample to those aged 40 or above because it is suggested that ageing is a non-linear process with first substantial acceleration in the 40s (Shen et al., 2024). All analyses were implemented in Stata 18.

## Results

### Descriptives

The average age of our analytical sample was 52.3 years compared to 47.5 years in the original sample, and there was an underrepresentation of single persons (10.5% vs 22.5%). However, the analytical sample was reasonably evenly distributed across household income quintiles, and the distributions of other demographic, socioeconomic, behavioural and health factors were largely similar to the original sample (Table 1).

**Table 1.** Analytical sample characteristics compared to the original wave 2 sample.

| Variables |  | Analytical sample<br>(n=3,324)<br>%/mean (SD) | Original sample<br>(n= 46,891)<br>%/mean (SD)<sup>†</sup> |
| --- | --- | --- | --- |
| Sex: | Male | 44.2% | 48.3% |
|  | Female | 55.8% | 51.7% |
| Age: |  | 52.3 (15.2) | 47.5 (18.9) |
| Living with children: | Yes | 28.1% | 30.3% |
|  | No | 71.9% | 69.7% |
| Marital status: | Single | 10.5% | 22.5% |
|  | Married/cohabitating | 73.6% | 63.7% |
|  | separated/divorced/widowed | 15.8% | 13.8% |
| Household income: | Q1-lowest | 17.6% | 19.4% |
|  | Q2 | 19.1% | 19.3% |
|  | Q3 | 20.8% | 19.3% |
|  | Q4 | 22.0% | 20.1% |
|  | Q5-highest | 20.5% | 21.9% |
| Area of living: | Urban | 71.5% | 75.9% |
|  | Rural | 28.5% | 24.1% |
| Education qualification: | No qualification | 12.6% | 14.6% |
|  | GCSE or below | 33.7% | 31.7% |
|  | A level or above | 32.4% | 32.4% |
|  | Degree or above | 21.3% | 21.4% |
| Employment status: | Employed | 58.5% | 57.6% |
|  | Other | 41.5% | 42.4% |
| Area deprivation: | Q1-most deprived | 13.8% | 17.6% |
|  | Q2 | 18.1% | 18.8% |
|  | Q3 | 23.2% | 21.4% |
|  | Q4 | 22.1% | 21.1% |
|  | Q5-least deprived | 22.8% | 21.0% |
| Smoking status: | Never smoker | 40.7% | 41.3% |
|  | Ex-smoker | 41.1% | 37.3% |
|  | Current smoker | 18.2% | 21.4% |
| Drinking: |  | 4.3 (1.9) | 4.7 (2.2) |
| Long-standing illness: | Yes | 38.8% | 35.2% |
|  | No | 61.2% | 64.8% |

Notes: <sup>†</sup> Wave 2 cross-sectional weights were applied for the original sample

ACEng was relatively common among participants, with 82% of people doing three or more activities and 27.9% engaging in 11 or more activities (Table 2). More than three-quarters of people engaged in ACEng monthly or weekly.

**Table 2.** Descriptive statistics of the exposure and outcome variables (n=3,324)

| Variables | %/mean (SD) | Min-max |
| --- | --- | --- |
| ACEng frequency: ≤1/2 yearly | 10.3% | -- |
| 3+ yearly | 12.4% | -- |
| Monthly | 16.7% | -- |
| Weekly | 60.6% | -- |
| ACEng diversity: Low: 0-2 | 18.0% | -- |
| Medium: 3-6 | 30.1% | -- |
| High:7-10 | 24.1% | -- |
| Very high: 11+ | 27.9% | -- |
| PA frequency: No | 18.7% | -- |
| < monthly | 15.8% | -- |
| Monthly | 15.7% | -- |
| Weekly | 49.8% | -- |
| PA diversity: None | 19.3% | -- |
| Low: 1 | 20.3% | -- |
| Medium: 2-3 | 28.7% | -- |
| High: 4+ | 31.8% | -- |
| PA activeness: Not active | 23.5% | -- |
| Low | 18.7% | -- |
| Medium | 19.8% | -- |
| High | 19.6% | -- |
| Very high | 18.3% | -- |
| Hannum: | 50.7 (11.0) | 19.9-81.2 |
| Horvath2013: | 57.5 (10.4) | 26.6-87.5 |
| Horvath2018: | 50.6 (12.4) | 15.0-87.5 |
| Lin: | 46.3 (12.3) | 10.3-84.0 |
| PhenoAge: | 45.3 (12.3) | 8.2-82.5 |
| DunedinPoAm: | 1.0 (0.1) | 0.8-1.2 |
| DunedinPACE | 1.1 (0.1) | 0.7-1.5 |

PA had a relatively low diversity: 19.6% of participants did not do any PA, and less than a third engaged in four or more activities. However, PA frequency was higher, with nearly half of the participants engaging weekly. PA activeness was roughly evenly distributed in quintiles.

### ACEng and epigenetic ageing

For frequency of ACEng, there was no evidence of association with any of the first-generation clocks (Figure 1a). But associations were found with the second-generation PhenoAge clock and the third-generation DunedinPoAm and DunedinPACE clocks. For PhenoAge, although no evidence was found for the difference between the two low-frequency groups, epigenetic ageing was 0.77 years lower in people engaging monthly (95% CI=[-1.48, -0.05], p=0.036), and 0.79 years lower in people engaging weekly (95% CI=[-1.43, -0.14], p=0.017) compared to one or two times yearly. For DunedinPoAm, ACEng frequency of at least three times yearly (95% CI=[-0.02, -0.002], p=0.019), monthly (95% CI=[-0.02, -0.006], p=0.001) and weekly (95% CI=[-0.01, -0.0004], p=0.037) were associated with a slower pace of epigenetic ageing by 0.01 biological years per one chronological year. The DunedinPACE coefficients were larger in magnitude than those for DunedinPoAm. Engagement at least three times yearly was associated with a slower pace of epigenetic ageing by 0.02 (95% CI=[-0.03, -0.003], p=0.107), although this association was not statistically significant. Engagement weekly (95% CI=[-0.04,-0.01], p=0.004) and monthly (95% CI=[-0.04,-0.01], p=0.002) were associated with a slower pace by 0.03.

**Figure 1.**
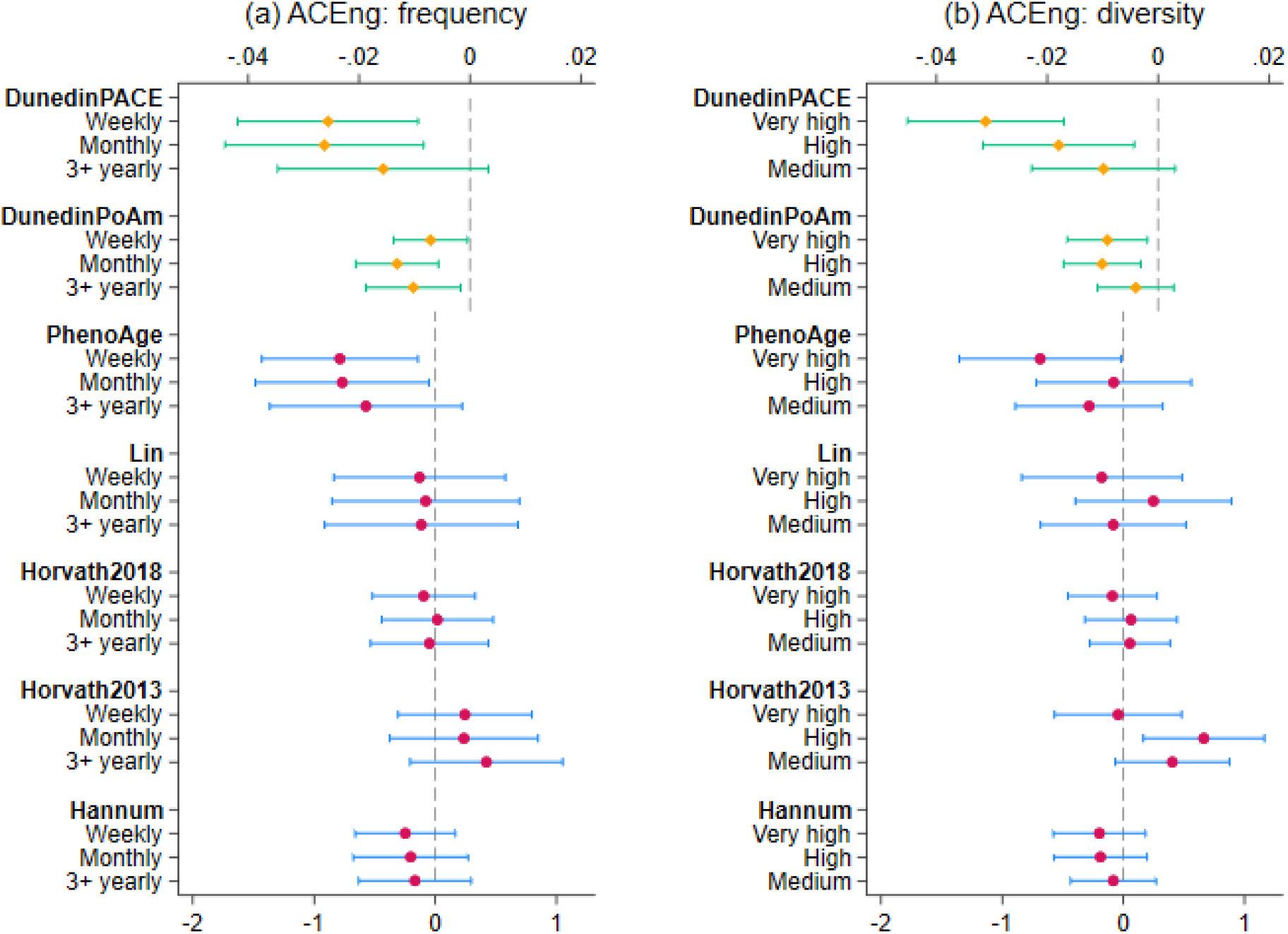
Estimated average treatment effect and 95% confidence intervals for ACEng diversity and frequency measures from doubly robust estimation using IPWRA (n=3,324)

For diversity of ACEng, there was again no evidence of association with any of the first-generation clocks. However, there was some evidence that the highest level of diversity was associated with a lower value of the second-generation PhenoAge clock (ATE=-0.69 years lower, 95%CI=[-1.35, -0.02], p=0.044) (Figure 1b). For the third-generation DunedinPoAm clock, there was no significant difference between medium and low diversity levels. However, both high diversity (95% CI=[-0.02, -0.003], p=0.005) and very high diversity (95% CI=[-0.02, -0.002], p=0.013) were associated with a slower pace of epigenetic ageing by 0.01. Similarly, for DunedinPACE, no significant difference was observed between medium and low diversity levels. However, high diversity was associated with a slower pace by 0.02 (95% CI=[-0.03, -0.004], p=0.01), and very high diversity by 0.03 (95% CI=[-0.05, -0.02], p<0.001).

### PA and epigenetic ageing

For frequency of PA, there was no evidence of association with any of the first- or second-generation clocks (Figure 2a). But for the DunedinPoAm clock, weekly engagement was associated with slower pace of epigenetic ageing by 0.01 (95% CI=[-0.02,-0.004], p=0.001). For DunedinPACE, less than monthly engagement was associated with a lower pace of epigenetic ageing by 0.02 (95% CI=[-0.03,-0.003], p=0.017), monthly engagement by 0.03 (95% CI=[-0.04,-0.01], p=0.001), and weekly engagement by 0.03 (95% CI=[-0.04,-0.02], p<0.001).

**Figure 2.**
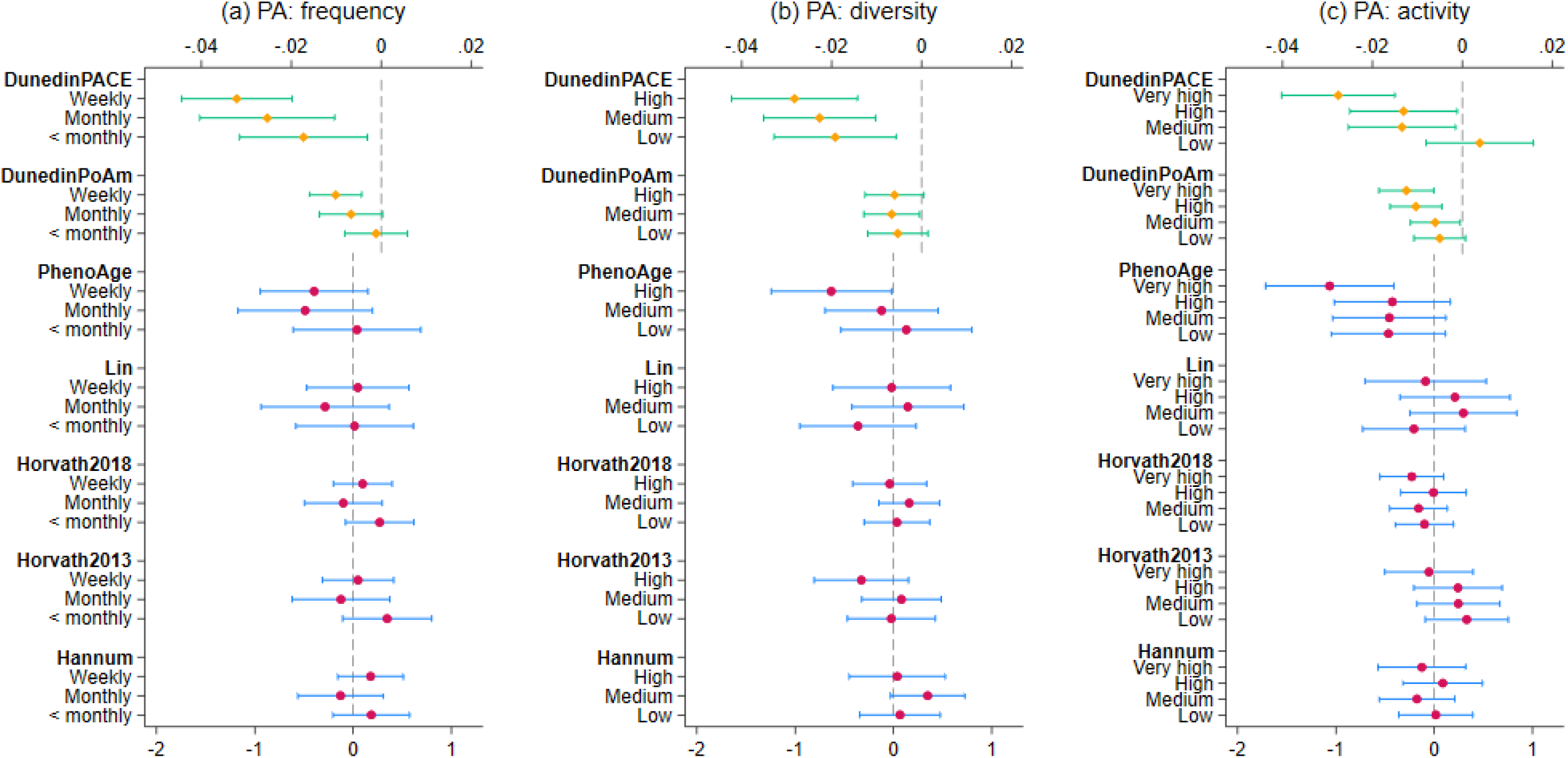
Estimated average treatment effect and 95% confidence intervals for PA diversity, frequency and activeness from doubly robust estimation using IPWRA (n=3,324)

For diversity of PA, there was again no evidence of association with any of the first-generation clocks (Figure 2b). Some evidence was found for the PhenoAge clock: the highest level of diversity was associated with a lower epigenetic ageing by 0.63 years (XX months; 95% CI=[-1.24, -0.02], p=0.043). For DunedinPoAm, medium diversity was associated with a slower pace of epigenetic ageing by 0.007 (95% CI=[-0.01, -0.0005], p=0.035) compared to no PA. Most consistent evidence was found for DunedinPACE where low (ATE=-0.02, 95% CI=[-0.03,-0.01], p=0.006), medium (ATE=-0.02, 95% CI=[-0.04,-0.01], p<0.001) and high (ATE=-0.03, 95% CI=[-0.04,-0.01], p<0.001) diversity were all associated with a slower pace of epigenetic ageing.

### Sensitivity analyses

There was no evidence of associations in first-generation clocks for levels of activeness (Figure 2c). However, very high levels of PA activeness were associated with 1.06 years lower PhenoAge (95% CI=[-1.72, -0.41], p=0.001). And for DunedinPoAm, medium levels of activeness was associated with a lower pace of epigenetic ageing by -0.006 (95% CI=[-0.01, -0.0005], p=0.033), high levels of activeness by 0.01 (95% CI=[-0.02, -0.005], p<0.001), and very high levels of activeness by 0.01 (95% CI=[-0.02, -0.006], p<0.001.For DunedinPACE, medium (95% CI=[-0.03,-0.001], p=0.028) and high (95% CI=[-0.03,-0.001], p=0.032) levels of activeness were associated with a lower pace of epigenetic ageing by 0.01, and very high levels of activeness by 0.03 (95% CI=[-0.04,-0.01], p<0.001)

The associations of ACEng and PA with PhenoAge, DunedinPoAm and DunedinPACE persisted even after accounting for BMI (Figure S2 & S3 in the Supplement). The results from sensitivity analyses restricting the sample to those aged 40 or above were consistent with the main results, with generally larger effect sizes (Figure S4 & S5 in the Supplement).

## Discussion

Using an outcome-wide approach involving seven epigenetic clocks, we found associations between two health-promoting leisure activities – arts and cultural engagement (ACEng) and physical activity (PA) – and slower epigenetic ageing. Specifically, ACEng and PA were related to PhenoAge, DunedinPoAm and DunedinPACE clocks, although not to the other measured clocks, with comparable effect sizes between ACEng and PA (Lin, Horvath2018, Horvath2013 and Hannum). For ACEng, diversity and frequency of engagement were related to DunedinPoAm and DunedinPACE, while for PhenoAge, there was a slightly clearer relationship for frequency than diversity. For PA, higher levels of frequency, diversity, and activeness were related to DunedinPoAm and DunedinPACE, while only the highest diversity and activeness were related to PhenoAge. These findings were all stronger amongst middle-aged and older adults.

This was the first study to show a relationship between ACEng and epigenetic ageing. It builds on strong theoretical and empirical underpinnings for why the arts could affect fundamental biological hallmarks of ageing. Life-course psychosocial stressors have been clearly linked with accelerated epigenetic ageing and broader physiological wear and tear across tissues and organ systems (Zannas, 2016). One of the fundamental mechanistic effects of arts engagement is reductions in psychophysiological markers of stress, demonstrated in clinical and non-clinical studies (de Witte et al., 2022; Finn & Fancourt, 2018; Lee et al., 2023). Notably, variety of engagement has been proposed as key here as it provides opportunities for diverse exposure to active ingredients, formation of multiple identities and even increased social capital (i.e. tangible and intangible resources), all of which support with buffering of stressors (Cohen & Wills, 1985). So, it is significant that variety and frequency were both related to slower epigenetic ageing. For both ACEng and PA, reductions in inflammatory pathways (which are well-reported for both) (Fancourt, Perkins, et al., 2016; Fancourt, Williamon, et al., 2016; Silverman & Deuster, 2014)may also be important mechanisms between engagement and epigenetic alterations. Anti-inflammatory effects of ACEng and PA engagement have been linked to methylation status as well as being a hallmark of ageing (“inflammageing”) (Zhu et al., 2021). Additionally, improvements in cardiovascular risk have been demonstrated to be mediators of the link between PA and epigenetic ageing (Fox et al., 2023), and this may also be the case for ACEng, for which there is strong mechanistic evidence of benefits for diverse cardiometabolic traits (Cao & Zhang, 2023; McCrary & Altenmüller, 2021; Peng et al., 2020). Notably, the findings were independent of BMI, which is important given that BMI has been strongly linked to epigenetic age both observationally and experimentally (Galkin et al., 2023).

Notably, we only found results for so-called ‘second-generation’ and ‘third-generation’ clocks but not for ‘first-generation’ clocks. This echoes some previous studies. The Rhineland study found results for second-generation clocks (PhenoAge and GrimAge) but not for first-generation clocks (Horvath2013 and Hannum), and a study using The Irish Longitudinal Study of Ageing (TILDA) found associations between physical performance (walking speed) and second- but not first-generation clocks (Fox et al., 2023; McCrory et al., 2021). Null findings for first-generation clocks have previously been shown for measures related to physical performance in multiple previous studies (Maddock et al., 2020; Quach et al., 2017). There are several reasons why second- and third-generation clocks may be more relevant to picking up decelerated ageing associated with leisure engagement. First-generation clocks are generally less sensitive predictors of age-related decline in clinical health measures (Horvath & Raj, 2018). This is because they do not incorporate clinical biomarkers in their derivation and hence are less sensitive to capturing the epigenetic ageing deceleration that results from biobehavioural factors like protective health behaviours (McCrory et al., 2020). First-generation clocks were also trained on cross-sectional data, which, unlike longitudinal data, do not account for mortality selection. This biases the algorithm to select markers that are correlative with ageing rather than causal, because causal loci that should exhibit diminishing age prediction in later life as the individuals exhibiting these traits are progressively selected out of the cohort (Nelson et al., 2020). By using multiple different generations of ageing clocks, our study provides a clearer demonstration of the differential findings between earlier and more recent clocks.

Our study has many strengths, including using a representative cohort study, rich measures of both frequency and diversity of behaviours for our two leisure outcomes, adoption of an outcome-wide approach to epigenetic clocks, and consideration of diverse confounding factors. However, there are some limitations. First, we relied on participants’ self-report on their behaviours, which brings the risk of recall bias and self-report bias. However, some of our clocks overlapped with the Rhineland study, and were corroborated, which is reassuring given that study used objective assessments of behaviours. Second, we included all identified confounding factors, including a particular focus on diverse measures of SEP. But unidentified or unmeasured confounding remains a risk. Nonetheless, we adopted a doubly-robust estimation approach (a methodological advance on previous work relating lifestyle factors to epigenetic clocks), which allows for misspecification in confounders for either the exposure or the outcome. We relied on DNAm present in whole blood. However, the effect of leisure on the epigenome is likely not uniform across the body. This is particularly important for PA, where ageing deceleration may be different in, say, muscle tissues. So, future studies focusing on more specific DNAm tissue data are encouraged. Finally, while we have examined six different clocks within this set of analyses, this is not exhaustive. Other clocks may show different sets of findings.

Overall, our results provide the first evidence that ACEng is related to epigenetic ageing, suggesting the value of its exploration alongside other lifestyle factors (Galkin et al., 2023). In particular, diversity of engagement appears as important as frequency of engagement. It is also of note that the effect size was comparable for ACEng and for PA with respect to epigenetic ageing. ACEng is a much more recently recognised health behaviour, but these findings suggest the importance of continued exploration into its health effects alongside other more established health behaviours. We also extend the existing understanding of the relationship between PA and epigenetic ageing, demonstrating a clear association with second- but not first-generation epigenetic clocks and extending analyses to new clocks not included in previous analyses. Our findings are relevant for several reasons. First, decelerations in ageing clocks, including those within our study, have been demonstrated to have clinical (as well as statistical) importance, including improvements in physical performance, polypharmacy, cognitive state and all-cause mortality risk (McCrory et al., 2021). Indeed, our strongest results involved DunedinPACE, which has shown improved performance than DunedinPoAm and has been related to improved performance in physical and physiological measures of ageing over subsequent years (Belsky et al., 2022). And it is notable that associations between leisure and epigenetic ageing became more prominent in adults from mid-life. Second, recent work suggests that epigenetic ageing is potentially reversible (Fahy et al., 2019). The persistence of epigenetic changes in response to modifiable behaviours such as leisure engagement is greatly underexplored. However, given the experimental evidence reviewed earlier on the effects of both ACEng and PA on DNAm generally and epigenetic clocks specifically (Fiorito et al., 2021; Kanduri, Raijas, et al., 2015; Nair et al., 2021), future intervention studies could explore whether lifestyle changes have any value to slowing or potentially reversing epigenetic ageing.

## Data Availability

Data are publicly available through UKDS https://beta.ukdataservice.ac.uk/datacatalogue/series/series?id=2000053

## Conflict of Interest

The authors report no conflict of interest.

## Data availability

Data of this study are publicly available via the UK Data service: https://ukdataservice.ac.uk/

## Funding

This study was supported by a Wellcome Discovery Award [326117/Z/25/Z] and The EpiArts Lab, a National Endowment for the Arts Research Lab at the University of Florida, is supported in part by an award from the National Endowment for the Arts (1936473-38-24). The opinions expressed are those of the authors and do not represent the views of the National Endowment for the Arts Office of Research & Analysis or the National Endowment for the Arts. The National Endowment for the Arts does not guarantee the accuracy or completeness of the information included in this material and is not responsible for any consequences of its use. The EpiArts Lab is also supported by Americans for the Arts, Bloomberg Philanthropies (F024567), and the Pabst Steinmetz Foundation. This paper received additional support from the UK Research and Innovation [MR/Y01068X/1]. LM was supported by a PhD studentship provided through the Soc-B Centre for Doctoral Training (CDT), funded by the Economic and Social Research Council (ESRC) and the Biotechnology & Biological Sciences Research Council (BBSRC). SF’s funding was provided as part of the Understanding Society fellowship programme, a component of the Study’s Economic and Social Research Council award [ES/S007253/1].

## Acknowledgements

FB acknowledges training received from the University of Michigan Genomics for Social Scientists Workshop (NIA R25 AG053227).

## Supplementary Materials

**Figure S1.**
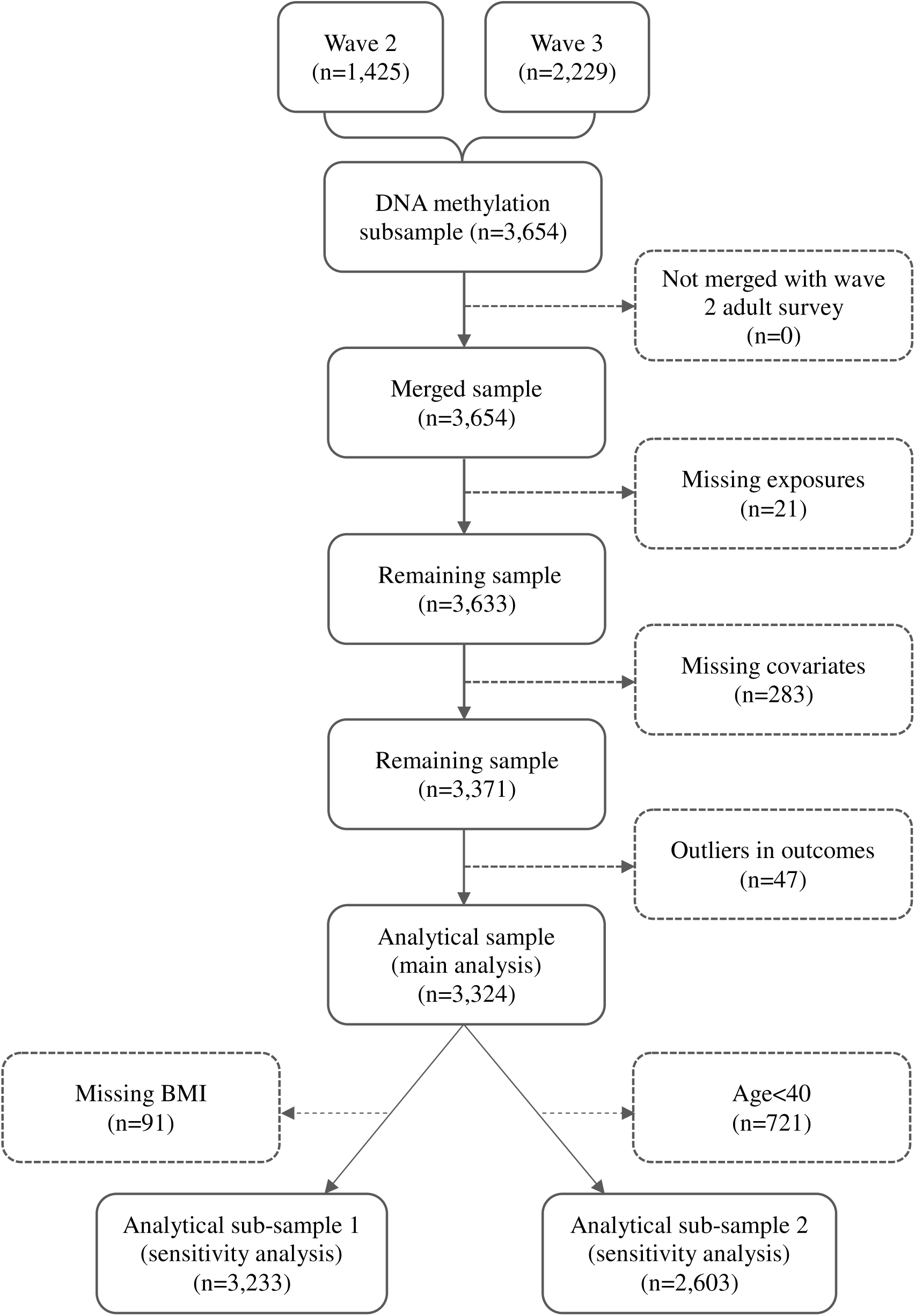
Sample selection diagram

**Figure S2.**
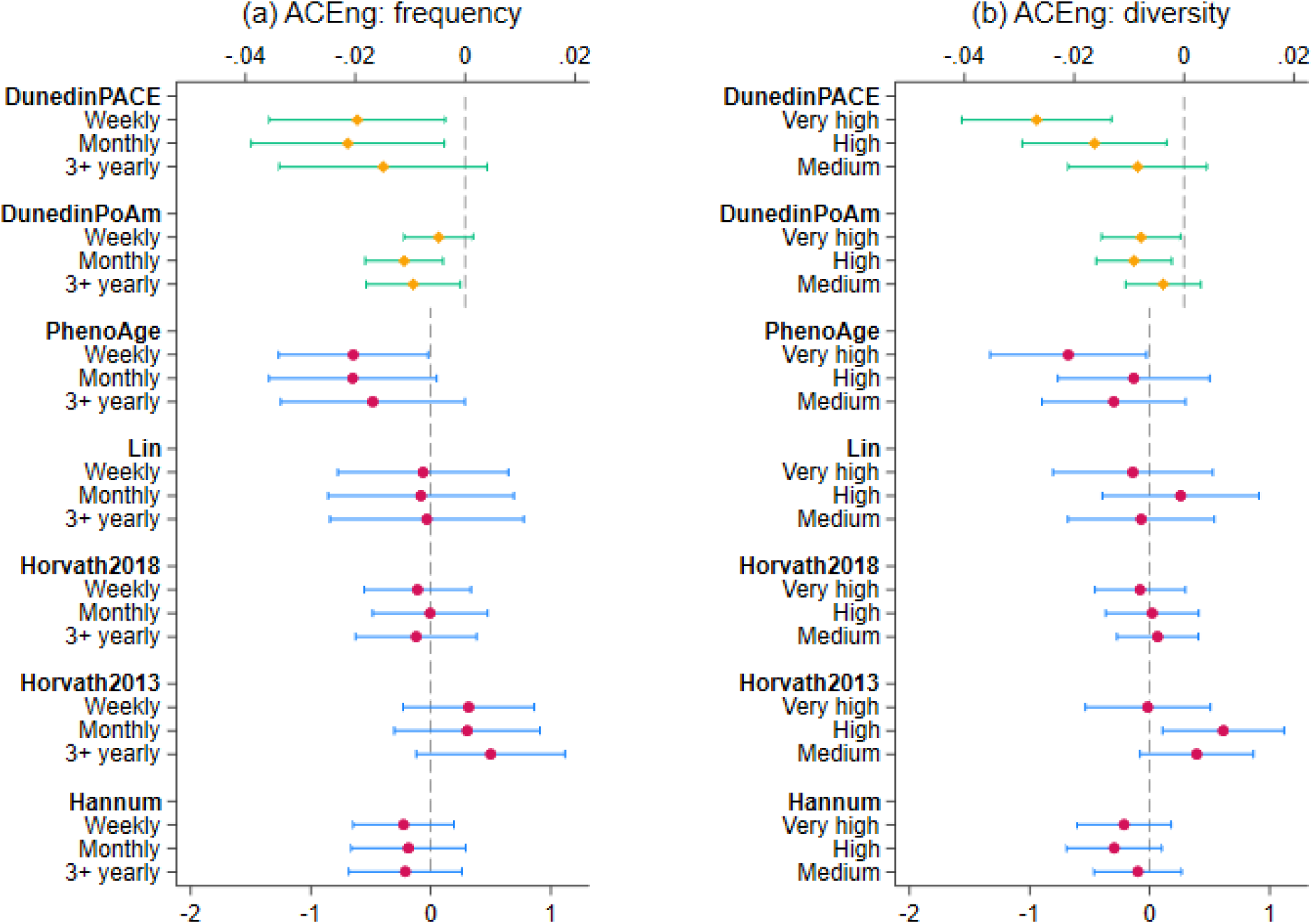
Estimated average treatment effect and 95% confidence intervals for ACEng diversity and frequency measures from doubly robust estimation using IPWRA (additionally controlling for BMI, n=3,233)

**Figure S3.**
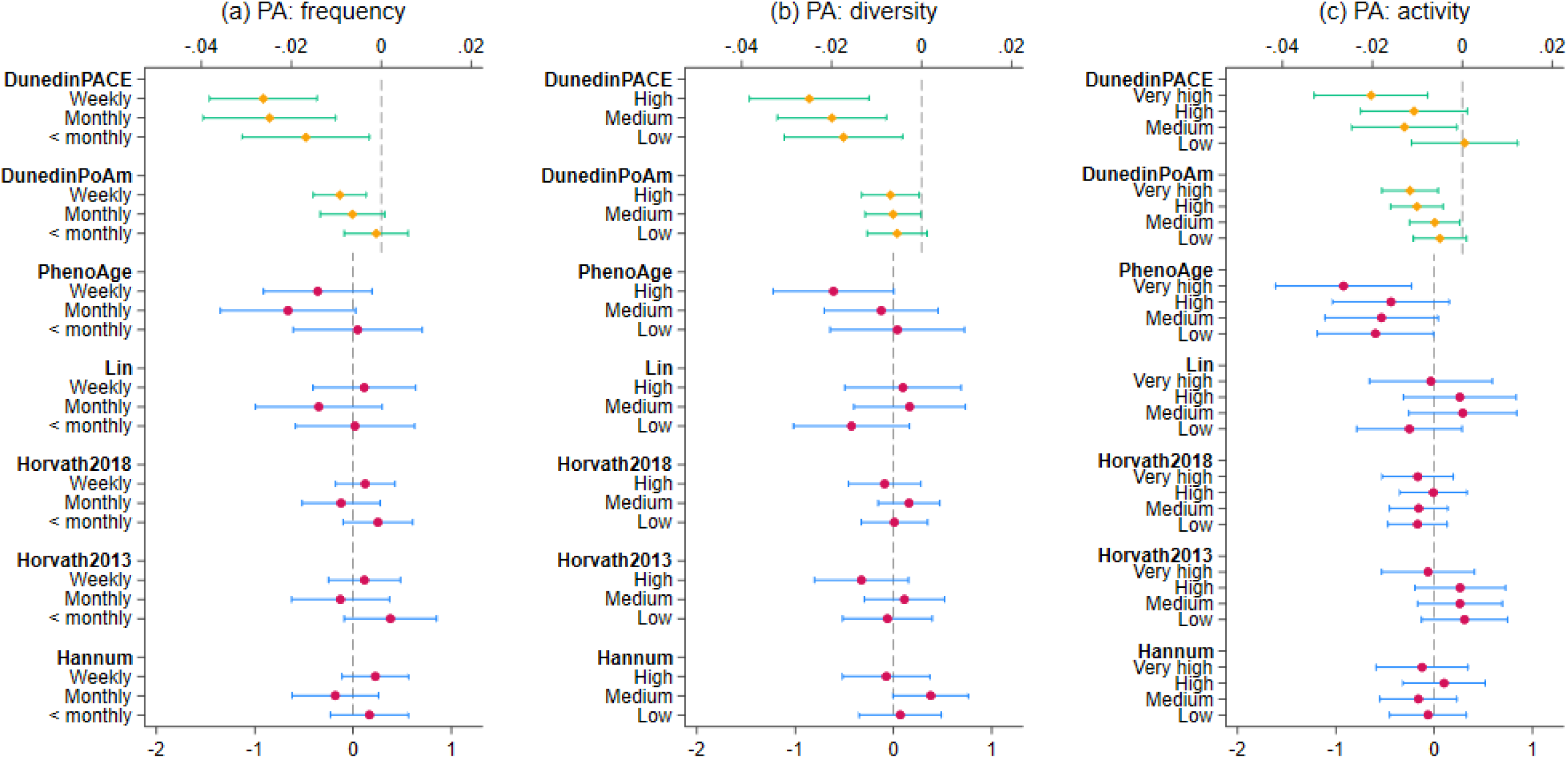
Estimated average treatment effect and 95% confidence intervals for PA diversity, frequency and activeness from doubly robust estimation using IPWRA (additionally controlling for BMI, n=3,233)

**Figure S4.**
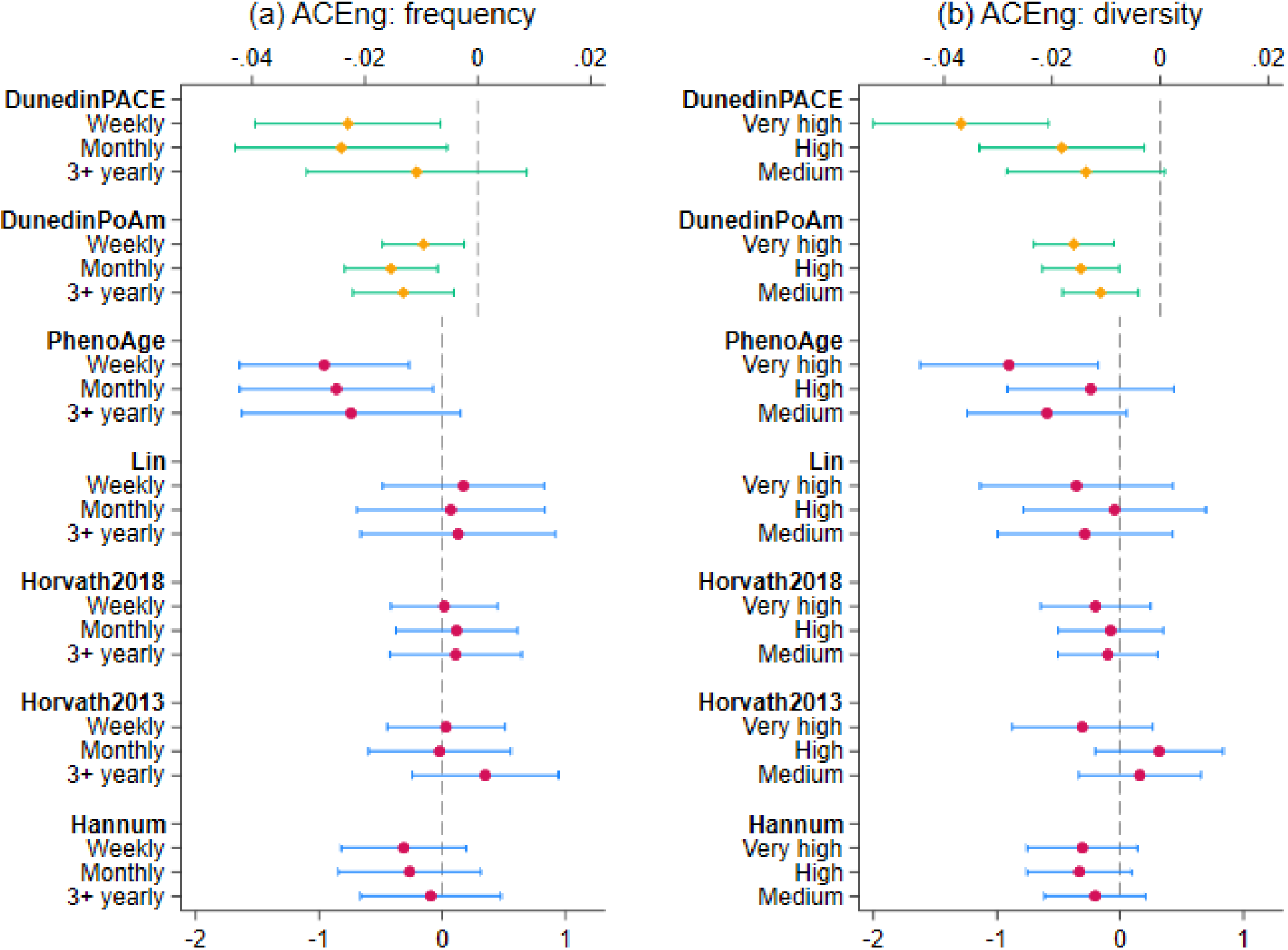
Estimated average treatment effect and 95% confidence intervals for ACEng diversity and frequency measures from doubly robust estimation using IPWRA (age>=40, n=2,603)

**Figure S5.**
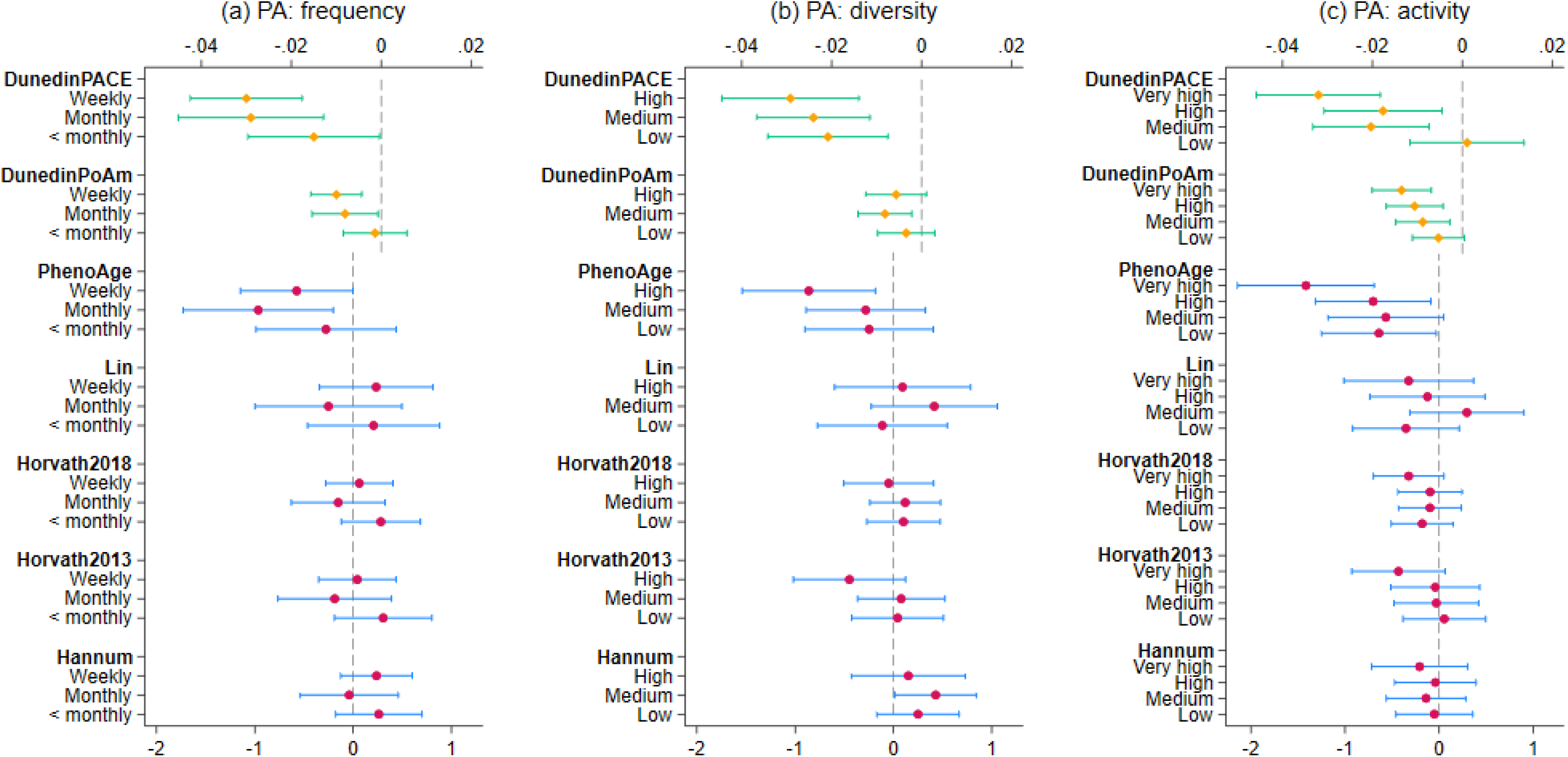
Estimated average treatment effect and 95% confidence intervals for PA diversity, frequency and activeness from doubly robust estimation using IPWRA (age>=40, n=2,603)

## Notes

### Competing Interest Statement

The authors have declared no competing interest.

### Funding Statement

This paper is supported by the UK Research and Innovation [MR/Y01068X/1]. LM was supported by a PhD studentship provided through the Soc-B Centre for Doctoral Training (CDT), funded by the Economic and Social Research Council (ESRC) and the Biotechnology & Biological Sciences Research Council (BBSRC)

### Author Declarations

All data were openly available prior to analyses commencing. Data can be accessed through https://www.understandingsociety.ac.uk/ and https://beta.ukdataservice.ac.uk/datacatalogue/series/series?id=2000053

### Summary of Updates

The previous funding statement accidentally omitted funders. This has been rectified.

## References

Bao, Y., Gorrie-Stone, T., & Kumari, M. (2022). Understanding Society: Waves 2-3 Nurse Health Assessment ‘Epigenetic Clocks’ derived from DNA methylation, 2010-2012.

Beard, J. R., & Bloom, D. E. (2015). Towards a comprehensive public health response to population ageing. The Lancet, 385(9968), 658–661. 10.1016/S0140-6736(14)61461-6

Bell, C. G., Lowe, R., Adams, P. D., Baccarelli, A. A., Beck, S., Bell, J. T., Christensen, B. C., Gladyshev, V. N., Heijmans, B. T., Horvath, S., Ideker, T., Issa, J. P. J., Kelsey, K. T., Marioni, R. E., Reik, W., Relton, C. L., Schalkwyk, L. C., Teschendorff, A. E., Wagner, W., … Rakyan, V. K. (2019). DNA methylation aging clocks: challenges and recommendations. Genome Biology 2019 20:1, 20(1), 1–24. 10.1186/S13059-019-1824-Y

Belsky, D. W., Caspi, A., Arseneault, L., Baccarelli, A., Corcoran, D., Gao, X., Hannon, E., Harrington, H. L., Rasmussen, L. J. H., Houts, R., Huffman, K., Kraus, W. E., Kwon, D., Mill, J., Pieper, C. F., Prinz, J., Poulton, R., Schwartz, J., Sugden, K., … Moffitt, T. E. (2020). Quantification of the pace of biological aging in humans through a blood test, the DunedinPoAm DNA methylation algorithm. ELife, 9, 1–56. 10.7554/ELIFE.54870

Belsky, D. W., Caspi, A., Corcoran, D. L., Sugden, K., Poulton, R., Arseneault, L., Baccarelli, A., Chamarti, K., Gao, X., Hannon, E., Harrington, H. L., Houts, R., Kothari, M., Kwon, D., Mill, J., Schwartz, J., Vokonas, P., Wang, C., Williams, B., & Moffitt, T. E. (2022). DunedinPACE, A DNA methylation biomarker of the Pace of Aging. ELife, 11. 10.7554/ELIFE.73420

Bittman, B., Berk, L., Shannon, M., Sharaf, M., Westengard, J., Guegler, K. J., & Ruff, D. W. (2005). Recreational music-making modulates the human stress response: A preliminary individualized gene expression strategy. Medical Science Monitor, 11(2), BR31–BR40. https://experts.llu.edu/en/publications/recreational-music-making-modulates-the-human-stress-response-a-p-2

Cao, M., & Zhang, Z. (2023). Adjuvant music therapy for patients with hypertension: a meta-analysis and systematic review. BMC Complementary Medicine and Therapies, 23(1), 1–11. 10.1186/S12906-023-03929-6/FIGURES/7

Cohen, S., & Wills, T. A. (1985). Stress, Social Support, and the Buffering Hypothesis. Psychological Bulletin, 98(2), 310–357. 10.1037/0033-2909.98.2.310

Crimmins, E. M., Klopack, E. T., & Kim, J. K. (2024). Generations of epigenetic clocks and their links to socioeconomic status in the Health and Retirement Study. Epigenomics. 10.1080/17501911.2024.2373682

de Witte, M., Pinho, A. da S., Stams, G. J., Moonen, X., Bos, A. E. R., & van Hooren, S. (2022). Music therapy for stress reduction: a systematic review and meta-analysis. Health Psychology Review, 16(1), 134–159. 10.1080/17437199.2020.1846580

Fahy, G. M., Brooke, R. T., Watson, J. P., Good, Z., Vasanawala, S. S., Maecker, H., Leipold, M. D., Lin, D. T. S., Kobor, M. S., & Horvath, S. (2019). Reversal of epigenetic aging and immunosenescent trends in humans. Aging Cell, 18(6), e13028. 10.1111/ACEL.13028

Fancourt, D., Aughterson, H., Finn, S., Walker, E., & Steptoe, A. (2021). How leisure activities affect health: a narrative review and multi-level theoretical framework of mechanisms of action. The Lancet Psychiatry. 10.1016/S2215-0366(20)30384-9

Fancourt, D., & Finn, S. (2019). WHO Health Evidence Synthesis Report-Cultural Contexts of Health: The role of the arts in improving health and well-being in the WHO European Region.

Fancourt, D., Perkins, R., Ascenso, S., Atkins, L., Kilfeather, S., Carvalho, L., Steptoe, A., & Williamon, A. (2016). Group Drumming Modulates Cytokine Response in Mental Health Services Users: A Preliminary Study. Psychotherapy and Psychosomatics, 85(1), 53–55. 10.1159/000431257

Fancourt, D., Williamon, A., Carvalho, L. A., Steptoe, A., Dow, R., & Lewis, I. (2016). Singing modulates mood, stress, cortisol, cytokine and neuropeptide activity in cancer patients and carers. Ecancermedicalscience, 10, 631. 10.3332/ECANCER.2016.631

Finn, S., & Fancourt, D. (2018). The biological impact of listening to music in clinical and nonclinical settings: A systematic review. Progress in Brain Research, 237, 173–200. 10.1016/BS.PBR.2018.03.007

Fiorito, G., Caini, S., Palli, D., Bendinelli, B., Saieva, C., Ermini, I., Valentini, V., Assedi, M., Rizzolo, P., Ambrogetti, D., Ottini, L., & Masala, G. (2021). DNA methylation-based biomarkers of aging were slowed down in a two-year diet and physical activity intervention trial: the DAMA study. Aging Cell, 20(10), e13439. 10.1111/ACEL.13439;JOURNAL:JOURNAL:14749726;PAGE:STRING:ARTICLE/CHAPTER

Fox, F. A. U., Liu, D., Breteler, M. M. B., & Aziz, N. A. (2023). Physical activity is associated with slower epigenetic ageing—Findings from the Rhineland study. Aging Cell, 22(6), e13828. 10.1111/ACEL.13828;PAGEGROUP:STRING:PUBLICATION

Galkin, F., Kovalchuk, O., Koldasbayeva, D., Zhavoronkov, A., & Bischof, E. (2023). Stress, diet, exercise: Common environmental factors and their impact on epigenetic age. Ageing Research Reviews, 88, 101956. 10.1016/J.ARR.2023.101956

Hogg, M. A. (2016). Social Identity Theory. 3–17. 10.1007/978-3-319-29869-6_1

Horvath, S. (2013). DNA methylation age of human tissues and cell types. Genome Biology, 14(10), 1–20. 10.1186/GB-2013-14-10-R115/FIGURES/8

Horvath, S., Oshima, J., Martin, G. M., Lu, A. T., Quach, A., Cohen, H., Felton, S., Matsuyama, M., Lowe, D., Kabacik, S., Wilson, J. G., Reiner, A. P., Maierhofer, A., Flunkert, J., Aviv, A., Hou, L., Baccarelli, A. A., Li, Y., Stewart, J. D., … Raj, K. (2018). Epigenetic clock for skin and blood cells applied to Hutchinson Gilford Progeria Syndrome and ex vivo studies. Aging (Albany NY), 10(7), 1758. 10.18632/AGING.101508

Horvath, S., & Raj, K. (2018). DNA methylation-based biomarkers and the epigenetic clock theory of ageing. Nature Reviews Genetics 2018 19:6, 19(6), 371–384. 10.1038/s41576-018-0004-3

Jokai, M., Torma, F., McGreevy, K. M., Koltai, E., Bori, Z., Babszki, G., Bakonyi, P., Gombos, Z., Gyorgy, B., Aczel, D., Toth, L., Osvath, P., Fridvalszky, M., Teglas, T., Posa, A., Kujach, S., Olek, R., Kawamura, T., Seki, Y., … Radak, Z. (2023). DNA methylation clock DNAmFitAge shows regular exercise is associated with slower aging and systemic adaptation. GeroScience, 45(5), 2805–2817. 10.1007/S11357-023-00826-1/FIGURES/6

Kanduri, C., Kuusi, T., Ahvenainen, M., Philips, A. K., Lähdesmäki, H., & Järvelä, I. (2015). The effect of music performance on the transcriptome of professional musicians. Scientific Reports, 5(1), 1–7. 10.1038/SREP09506;TECHMETA=38,61;SUBJMETA=114,2019,208,212,2407,631;KWRD=MICROARRAYS,TRANSCRIPTOMICS

Kanduri, C., Raijas, P., Ahvenainen, M., Philips, A. K., Ukkola-Vuoti, L., Lähdesmäki, H., & Järvelä, I. (2015). The effect of listening to music on human transcriptome. PeerJ, 2015(3), e830. 10.7717/PEERJ.830/SUPP-6

Kresovich, J. K., Garval, E. L., Martinez Lopez, A. M., Xu, Z., Niehoff, N. M., White, A. J., Sandler, D. P., & Taylor, J. A. (2021). Associations of Body Composition and Physical Activity Level With Multiple Measures of Epigenetic Age Acceleration. American Journal of Epidemiology, 190(6), 984–993. 10.1093/AJE/KWAA251

Lee, H. Y., Nam, E. S., Chai, G. J., & Kim, D. M. (2023). Benefits of Music Intervention on Anxiety, Pain, and Physiologic Response in Adults Undergoing Surgery: A Systematic Review and Meta-analysis. Asian Nursing Research, 17(3), 138–149. 10.1016/J.ANR.2023.05.002

Levine, M. E., Lu, A. T., Quach, A., Chen, B. H., Assimes, T. L., Bandinelli, S., Hou, L., Baccarelli, A. A., Stewart, J. D., Li, Y., Whitsel, E. A., Wilson, J. G., Reiner1, A. P., Aviv1, A., Lohman, K., Liu, Y., Ferrucci, L., & Horvath, S. (2018). An epigenetic biomarker of aging for lifespan and healthspan. Aging (Albany NY), 10(4), 573. 10.18632/AGING.101414

Lin, Q., & Wagner, W. (2015). Epigenetic Aging Signatures Are Coherently Modified in Cancer. PLOS Genetics, 11(6), e1005334. 10.1371/JOURNAL.PGEN.1005334

López-Otín, C., Blasco, M. A., Partridge, L., Serrano, M., & Kroemer, G. (2023). Hallmarks of aging: An expanding universe. Cell, 186(2), 243–278. 10.1016/J.CELL.2022.11.001

Lynn, P. (2009). Sample Design for Understanding Society (2009–01). https://www.researchgate.net/publication/254412141

Maddock, J., Castillo-Fernandez, J., Wong, A., Cooper, R., Richards, M., Ong, K. K., Ploubidis, G. B., Goodman, A., Kuh, D., Bell, J. T., & Hardy, R. (2020). DNA Methylation Age and Physical and Cognitive Aging. The Journals of Gerontology: Series A, 75(3), 504–511. 10.1093/GERONA/GLZ246

McCrary, J. M., & Altenmüller, E. (2021). Mechanisms of Music Impact: Autonomic Tone and the Physical Activity Roadmap to Advancing Understanding and Evidence-Based Policy. Frontiers in Psychology, 12, 727231. 10.3389/FPSYG.2021.727231/BIBTEX

McCrory, C., Fiorito, G., Hernandez, B., Polidoro, S., O’Halloran, A. M., Hever, A., Ni Cheallaigh, C., Lu, A. T., Horvath, S., Vineis, P., & Kenny, R. A. (2021). GrimAge Outperforms Other Epigenetic Clocks in the Prediction of Age-Related Clinical Phenotypes and All-Cause Mortality. The Journals of Gerontology: Series A, 76(5), 741–749. 10.1093/GERONA/GLAA286

McCrory, C., Fiorito, G., McLoughlin, S., Polidoro, S., Cheallaigh, C. N., Bourke, N., Karisola, P., Alenius, H., Vineis, P., Layte, R., & Kenny, R. A. (2020). Epigenetic Clocks and Allostatic Load Reveal Potential Sex-Specific Drivers of Biological Aging. The Journals of Gerontology: Series A, 75(3), 495–503. 10.1093/GERONA/GLZ241

Nair, P. S., Kuusi, T., Ahvenainen, M., Philips, A. K., & Järvelä, I. (2019). Music-performance regulates micrornas in professional musicians. PeerJ, 2019(3), e6660. 10.7717/PEERJ.6660/SUPP-5

Nair, P. S., Raijas, P., Ahvenainen, M., Philips, A. K., Ukkola-Vuoti, L., & Järvelä, I. (2021). Music-listening regulates human microRNA expression. Epigenetics, 16(5), 554–566. 10.1080/15592294.2020.1809853;JOURNAL:JOURNAL:KEPI20;WGROUP:STRING:PUBLICATION

Nelson, P. G., Promislow, D. E. L., & Masel, J. (2020). Biomarkers for Aging Identified in Cross-sectional Studies Tend to Be Non-causative. The Journals of Gerontology: Series A, 75(3), 466–472. 10.1093/GERONA/GLZ174

Noroozi, R., Rudnicka, J., Pisarek, A., Wysocka, B., Masny, A., Boroń, M., Migacz-Gruszka, K., Pruszkowska-Przybylska, P., Kobus, M., Lisman, D., Zielińska, G., Iljin, A., Wiktorska, J. A., Michalczyk, M., Kaczka, P., Krzysztofik, M., Sitek, A., Ossowski, A., Spólnicka, M., … Pośpiech, E. (2024). Analysis of epigenetic clocks links yoga, sleep, education, reduced meat intake, coffee, and a SOCS2 gene variant to slower epigenetic aging. GeroScience, 46(2), 2583–2604. 10.1007/S11357-023-01029-4/TABLES/1

Pagiatakis, C., Musolino, E., Gornati, R., Bernardini, G., & Papait, R. (2021). Epigenetics of aging and disease: a brief overview. Aging Clinical and Experimental Research, 33(4), 737–745. 10.1007/S40520-019-01430-0/FIGURES/3

Peng, Y., Su, Y., Wang, Y. Di, Yuan, L. R., Wang, R., & Dai, J. S. (2020). Effects of regular dance therapy intervention on blood pressure in hypertension individuals: a systematic review and meta-analysis. The Journal of Sports Medicine and Physical Fitness, 61(2), 301–309. 10.23736/S0022-4707.20.11088-0

Quach, A., Levine, M. E., Tanaka, T., Lu, A. T., Chen, B. H., Ferrucci, L., Ritz, B., Bandinelli, S., Neuhouser, M. L., Beasley, J. M., Snetselaar, L., Wallace, R. B., Tsao, P. S., Absher, D., Assimes, T. L., Stewart, J. D., Li, Y., Hou, L., Baccarelli, A. A., … Horvath, S. (2017). Epigenetic clock analysis of diet, exercise, education, and lifestyle factors. Aging (Albany NY), 9(2), 419. 10.18632/AGING.101168

Sailani, M. R., Halling, J. F., Møller, H. D., Lee, H., Plomgaard, P., Pilegaard, H., Snyder, M. P., & Regenberg, B. (2019). Lifelong physical activity is associated with promoter hypomethylation of genes involved in metabolism, myogenesis, contractile properties and oxidative stress resistance in aged human skeletal muscle. Scientific Reports 2019 9:1, 9(1), 1–11. 10.1038/s41598-018-37895-8

Shen, X., Wang, C., Zhou, X., Zhou, W., Hornburg, D., Wu, S., & Snyder, M. P. (2024). Nonlinear dynamics of multi-omics profiles during human aging. Nature Aging 2024, 1–16. 10.1038/s43587-024-00692-2

Sillanpää, E., Ollikainen, M., Kaprio, J., Wang, X., Leskinen, T., Kujala, U. M., & Törmäkangas, T. (2019). Leisure-time physical activity and DNA methylation age - A twin study. Clinical Epigenetics, 11(1), 1–8. 10.1186/S13148-019-0613-5/FIGURES/1

Silverman, M. N., & Deuster, P. A. (2014). Biological mechanisms underlying the role of physical fitness in health and resilience. Interface Focus, 4(5). 10.1098/RSFS.2014.0040;WEBSITE:WEBSITE:RSJ-SITE;WGROUP:STRING:PUBLICATION

The Lancet Public Health. (2017). Ageing: a 21st century public health challenge? The Lancet Public Health, 2(7), e297. 10.1016/S2468-2667(17)30125-1,

Warran, K., Burton, A., & Fancourt, D. (2022). What are the active ingredients of ‘arts in health’ activities? Development of the INgredients iN ArTs in hEalth (INNATE) Framework. Wellcome Open Research 2022 7:10, 7, 10. 10.12688/wellcomeopenres.17414.2

Wooldridge, J. M. (2010). Econometric Analysis of Cross Section and Panel Data (2nd ed.). MIT Press.

Zannas, A. S. (2016). Editorial Perspective: Psychological stress and epigenetic aging - What can we learn and how can we prevent? Journal of Child Psychology and Psychiatry and Allied Disciplines, 57(6), 674–675. 10.1111/JCPP.12535;JOURNAL:JOURNAL:14697610;REQUESTEDJOURNAL:JOURNAL:14697610;WGROUP:STRING:PUBLICATION

Zhang, W., Qu, J., Liu, G. H., & Belmonte, J. C. I. (2020). The ageing epigenome and its rejuvenation. Nature Reviews Molecular Cell Biology, 21(3), 137–150. 10.1038/S41580-019-0204-5;SUBJMETA=136,176,208,631,7;KWRD=AGEING,EPIGENETICS

Zhu, X., Chen, Z., Shen, W., Huang, G., Sedivy, J. M., Wang, H., & Ju, Z. (2021). Inflammation, epigenetics, and metabolism converge to cell senescence and ageing: the regulation and intervention. Signal Transduction and Targeted Therapy 2021 6:1, 6(1), 1–29. 10.1038/s41392-021-00646-9

